# Association of coronary artery bypass with cognitive impairment in coronary artery disease across APO (ε) genotypes in AllofUS

**DOI:** 10.64898/2026.04.12.26350734

**Authors:** Praveen Hariharan, Minoo Bagheri, Emelia Asamoah, Ioana Voiculescu, Purnima Singh, Tafadzwa Machipisa, Tess Pottinger, Antone Opekun

## Abstract

**BACKGROUND:** Coronary artery bypass graft (CABG) is a widely performed procedure for coronary artery disease (CAD), yet its association with Impaired Cognition (IC), i.e., mild-cognitive impairment or all-cause dementia, while accounting for APO (ε) genotype, remains unclear.

**METHODS:** We analyzed AllofUS participants with CAD (Age≥60 yrs) from 2017-2023. We defined CAD as a history of angina/myocardial infarction/chronic ischemic heart disease or having percutaneous coronary intervention/CABG, and IC as mild cognitive impairment or all-cause dementia using ICD/SNOMED codes. We performed logistic regression analyses to assess the association between CABG and IC, adjusting for clinical factors (age, sex, hypertension, diabetes, hyperlipidemia, depression, stroke, smoking, alcohol use, statin/antihypertensive/antidiabetic use), social determinants (self-reported race/ethnicity, income, employment), and APO (ε) genotypes. We further performed stratified analyses across APO (ε) genotypes (ε2/ε2, ε2/ε3 ε3/ε3, ε2/ε4, ε3/ε4, ε4/ε4). We defined significance at p ≤ 0.05.

**RESULTS:** We included 22,349 with CAD and identified 908 with IC after CAD till 2023. 40% were females, 70% were White, 12% were Black, and 9% were Hispanic. The proportion of IC was higher (**5.1% vs 3.5%, p=1e-08**) in CABG (n=8,135) vs non-CABG (n=14,214). After adjusting for clinical factors, social determinants, and APO (ε) genotypes, CABG (**1.23;1.06-1.41, p = 0.005**) was associated with IC. In APO (ε) stratified analysis, the association of CABG with IC was strongest in the APO ε2/ε3 group (**1.91;1.21-3.02, p = 0.005**).

**CONCLUSION:** In the AllofUS cohort, we observed an association between CABG and IC in CAD participants, with the strongest association in the APO ε2/ε3 group.

**Key Message:** *What is already known on this topic:* Coronary artery disease (CAD) and Impaired Cognitive (IC) disease, i.e., mild cognitive impairment and all-cause dementia, share genetic, sociodemographic, and clinical factors, including cardiovascular conditions like coronary artery bypass grafting (CABG) procedure.

*What this study adds:* We observed an association between CABG and IC in CAD participants after adjusting for sociodemographic, clinical factors, and APO (ε) effects. Further, when CAD participants were stratified across APO (ε) groups, CABG was significantly associated with IC in the APO ε2/ε3 group.

*How this study might affect research, practice or policy:* Our observations highlight the role of APO (ε) genotype evaluation in CAD patients for IC risk assessment.

## BACKGROUND

Impaired Cognitive Disease (IC), i.e, all-cause dementia (ACD) or mild cognitive impairment (MCI), and Coronary Artery Disease (CAD) are a significant cause of morbidity in the US and globally, with associated increased financial burden to health care systems and patients.^1,2^ IC after a CAD event is a matter of concern and its prevalence increases with age (≥60 years).^1,3–6^

While IC as a clinical entity may have underlying heterogeneous neuropathologies, clinical factors like age, sex, diabetes, hypertension, hyperlipidemia, depression, ischemic stroke, smoking, alcohol use and certain cardiovascular conditions like coronary artery bypass grafting (CABG) are shared by CAD and IC patients.^2,7,8^ CABG is a widely performed surgical revascularization procedure for angiographically and clinically severe CAD, and studies evaluating association of CABG with IC have been limited due to small sample size, heterogenous outcome measures, and lack sex/racial/ethnic diversity.^5,9,10^ Further CABG as a procedure has been associated with IC through multiple biologic mechanisms including microemboli, neuronal apoptosis, neuroinflammation, blood brain barrier disruption, cerebral vascular dysregulation, and cerebral desaturation.^10^ In addition, more recently, through gene-based association (GBA) studies using large CAD biobanks, we identified many genes corresponding to neurodegenerative pathways to be significantly associated with CAD, underscoring the shared genetic underpinning of IC and CAD.^11^ In particular, we noted genes on chr 19.q13.32 (APOE) loci were significantly associated with CAD in the UK Biobank and CARDioGRAM cohort.^11^ Further, the APO (ε) genotype, defining single-nucleotide variants (SNVs, rs429359 and rs7412), demonstrates differential colocalization with other IC-associated SNVs that may occur based on the APO (ε) genotype.^12^ Also, APO (ε) genotype frequencies differ across ancestries, and APO (ε) genotype interacts differently across ancestries for various phenotype associations, including IC.^13^ APO (ε) genotypes have been identified to be jointly associated with CAD and IC, and can be a confounder when assessing the association of CABG with IC in CAD.^14–17^ Hence, the association of CABG with IC should not only be viewed as procedural outcomes but rather as CABG representing an IC risk factor milieu with underlying shared genotype substrates.^5^ However, large studies in CAD participants evaluating the association of CABG with IC while accounting for APO (ε) genotypes are lacking. Further, it is less clear if the association of CABG with IC varies across individual APO (ε) genotype groups in CAD in a diverse cohort. Understanding the association of CABG as a predictor for IC in CAD patients by having models accounting for APO (ε) genotypes would provide physicians with tools for heightened evaluation of CAD patients at risk of IC and offer opportunities for risk reduction.

Hence, we sought to evaluate the association of CABG with IC in all CAD participants enrolled in the diverse NIH AllofUS (AoU, v8) cohort between 2017 and 2023. We further assessed the association of CABG with IC across individual APO (ε) genotype groups.

## METHODS

### a) Study Design

**Figure 1** highlights the retrospective cohort design. We report our study according to STROBE guidelines.

**Figure 1:**
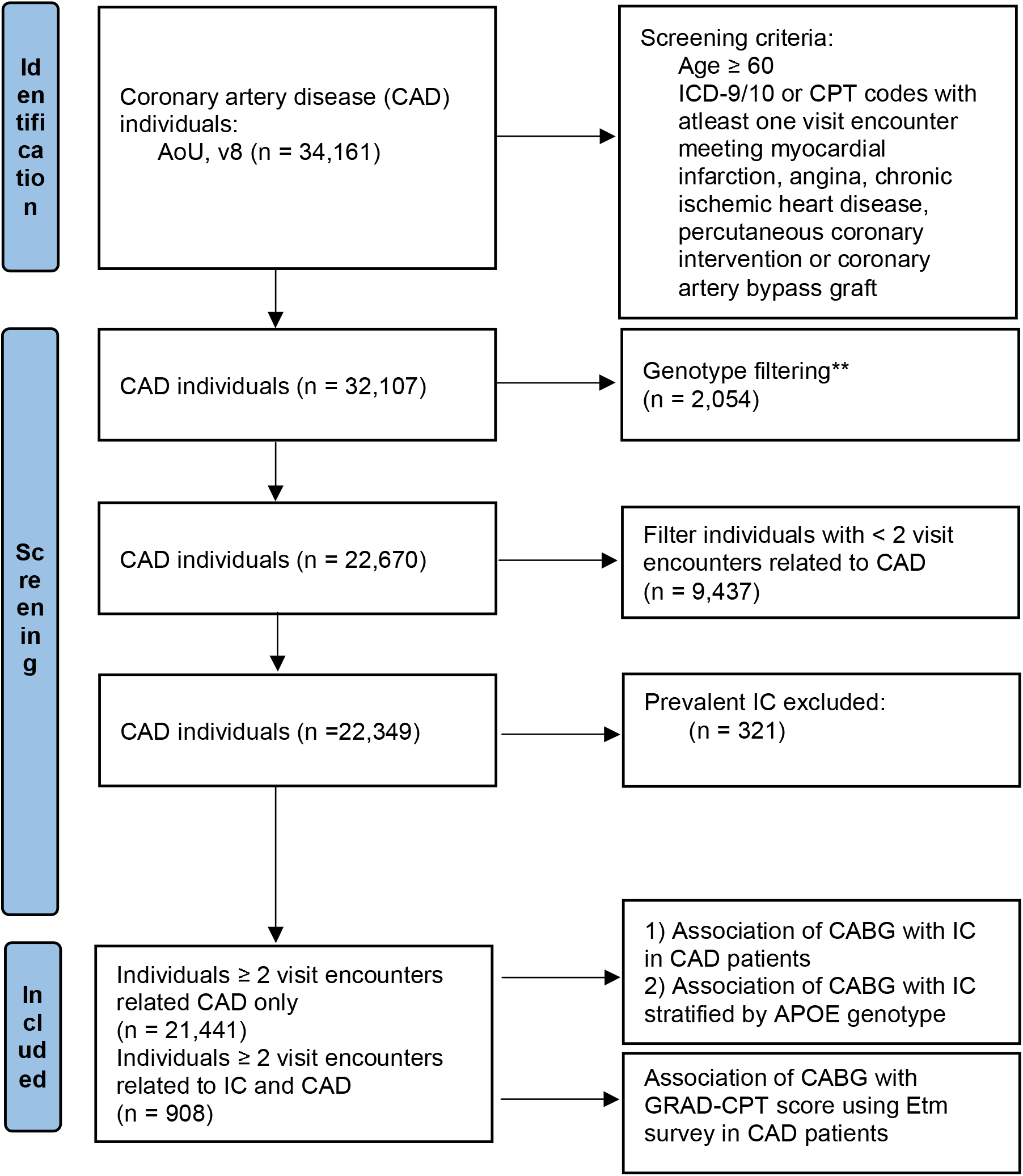
Study Design, *AllofUS* (AoU) Legend: CAD: Coronary Artery Disease IC: Impaired cognitive disease defined by individuals meeting ICD-9/10 for all cause dementia or mild cognitive impairment. **Genotype filtering: Removed related individuals (n=2,024), and performed sample and variant-level quality control with a call rate >95% (n=30).

### b) Setting and Data Sources

In brief, the AoU prospectively assembles diverse participants across 50 healthcare organizations in the US and has gathered surveys, electronic health records (EHR), biosamples, and genomic data since 2017.^18^ AoU utilizes the Observational Medical Outcomes Partnership Common Data Model (OMOP CDM) based on SNOMED (Systematized Nomenclature of Medicine), and/or ICD 9/10 codes to standardize data.^18^ We used the AoU version 8 (AoU, v8) dataset (C2024Q3R5, released Feb 2025) containing EHR data of more than 393,596 patients till 2023.^19^

### c) Selection of Cohort

We used OMOP CDM v 5.3.1 concept codes and names, which are based on SNOMED and ICD-9/10 codes to define CAD, IC, covariates, and used the associated index date of diagnoses (Details in Supplement 1).^4,20–22^

#### CAD definition

Any participant with 2 more visit encounters meeting one of the following criteria: A history of myocardial infarction (MI) (ICD-9: 410.x-412.x; ICD-10: I21.X-I24.X); the presence of stable or unstable angina (ICD-9: 413.x; ICD-10: I20.X); a history of chronic ischemic heart disease (ICD-9:414.x; ICD-10: I25.X); a history of percutaneous coronary intervention (PCI, ICD-10: Z95.5, Z98.61) or CABG (ICD-10: Z95.1).^4,11,23^

#### Primary outcome

We have defined IC when participants have 2 or more encounters meeting the ICD-9/10 diagnosis for all-cause dementia or mild cognitive impairment (290.X, 294.X, 331.X, F01.X, F02.X, F03.X, G30.X, G31.X).^4,20,24,25^ All events were captured via EHR till 2023 when AoU, v8 dataset was released. We defined IC to include the entire spectrum of cognitive impairment, required a minimum age of 50 years at IC diagnosis. We further excluded IC before CAD diagnosis to remove prevalent cases. The inclusion of age cut-offs and ICD codes has been shown to achieve good accuracy in defining IC in large registries.^20,25^

#### Secondary Outcome

To assess the association between CABG and neurocognitive test skills in CAD participants, we used the quantitative gradual onset continuous performance task (GradCPT) accuracy response (d-prime), collected using unsupervised Explore the Mind (EtM) surveys from AoU participants in 2023 (C2024Q3R5, released Feb 2025).^19^ GradCPT assesses core executive function by testing sustained attention and inhibitory control.^26^ We restricted our analysis to CAD participants who completed this optional EtM survey in 2023. Further, our primary and secondary outcomes definitions are consistent with the postoperative neurocognitive disorder (POCD) definition suggested by Evered et al.^27^

### d) Predictors

a. CABG: We defined CABG based on EHR diagnosis (Details in Supplement 1).
b. Clinical covariates: We further defined our covariates based on survey data and OMOP standard concept names and codes, which were based on SNOMED/ICD 9/10codes (Details in Supplement 1). 4,7,21,22

1. Demographics: Age, self-reported sex, and self-reported race/ethnicity.
2. Social determinants: We obtained the highest educational status, health insurance information, annual household income, and current employment status from baseline survey data received at the time of enrollment.
3. Clinical factors: Hypertension, diabetes, hyperlipidemia, ischemic stroke, depression, statin prescription data, antihypertensive prescription data (betablockers, angiotensin receptor blockers, calcium channel blockers, diuretics), antidiabetic prescription data (insulin, biguanides, sulfonylurea, thiazolidinediones, alpha glucosidase inhibitors, dipeptidyl peptidase 4 inhibitors, glucagon like peptide-1 inhibitors, sodium-glucose- cotransport 2 inhibitors) smoking, alcohol use, and Body Mass Index (BMI) data were obtained based on OMOP standard concept names and codes (Details in Supplement 1).^21,28^
4. Genotype covariates: We defined the APO (ε) genotype based on rs429358 (T/C) and rs7412 (C/T) single-nucleotide variants (SNV).^13^ For this analysis, we used the exon-joint call set with 45,704,594 variants across 414,797 individuals (AoU, version 8). We subsequently filtered the joint call set to include only our unique cases and controls. We further removed related samples and performed sample and variant-level QC with a call rate >95%. After filtering, we annotated rs429358 (T/C) and rs7412 (C/T) SNVs onto our CAD cohort. We further grouped APO (ε) into ε2/ε2 (rs429358:TT; rs7412:TT), ε2/ε3 (rs429358:TT; rs7412:CT), ε3/ε3 (rs429358:TT; rs7412:CC), ε2/ε4 (rs429358:CT; rs7412:CT), ε3/ε4 (rs429358:CT; rs7412:CC), ε4/ε4 (rs429358:CC; rs7412:CC) variant groups based on rs429358, rs7412 genotypes.^13^ Given ε3/ε3 has the most common genotype frequency, we used ε3/ε3 as the reference group for our analysis.^13^

### e) Standard Protocol Approvals, Registrations, and Patient Consents

Brown University Research Agreements and Contracting Committee reviewed our retrospective analysis proposal and approved the use of AoU researcher workbench. In addition, we have reviewed, completed AoU data access requirements and are in compliance with the data user code of conduct for registered and controlled tier data. There is no human/animal involved in this retrospective study.

### f) Statistical Analysis

#### 1) Primary Analyses

For our primary analyses, we included participants who met the CAD definition and had an age of≥ 60 for the final analysis. We subsequently queried for IC diagnoses in CAD participants and grouped participants into IC (with CAD) and CAD (without IC). We analyzed baseline characteristics across IC and CAD participants using simple means and proportions. We further compared the proportions of IC across all CAD participants with and without CABG. For univariate analysis, we used the chi-squared test. To assess association of CABG with IC in CAD participants, we performed logistic regression for CABG as independent variable with IC as dependent variable, while adjusting for clinical factors (age, sex, race/ethnicity, diabetes, hypertension, hyperlipidemia, depression, ischemic stroke, obesity (BMI ≥ 30), the smoking, alcohol use, statin use, antihypertensive use, antidiabetic use), social determinants (health insurance, education (college graduate and/or advanced degree), annual household income, employment status), and APO (ε) genotypes. We included self-reported race/ethnicity in the regression models to better adjust race and ethnicity as a social construct beyond APO (ε) genetics.^29^ We further repeated our regression models using genetically predicted ancestries (European, African, Admixed Americans, East Asian, South Asians, Middle Eastern).^18^ Subsequently, we assessed the association of CABG with IC in all CAD participants by running similar logistic regression models stratifying individuals by APO (ε) genotypes. For the EtM survey data analysis, we compared the GradCPT scores in CAD participants with and without CABG using a t-test. Finally, we assessed the association of CABG and the GradCPT scores using linear regression while adjusting for age, race/ethnicity, sex, stroke, alcohol use, smoking, health insurance, income, employment, educational status, and APO (ε) genotype. We defined significance at p < 0.05.

#### 2) Sensitivity Analysis

We performed the following sensitivity analyses to ensure consistency of our findings:

a. Propensity-score matched analyses: Given that the clinical factors in IC can confound the association of CABG between IC and CAD, we propensity-matched the IC (with CAD) to CAD (without IC) for many confounders (age, sex, race/ethnicity, diabetes, hypertension, stroke, depression, smoking, alcohol, hyperlipidemia, obesity, statin, antihypertensive use, antidiabetic use, education, income, employment and APO (ε) genotype). We ran similar logistic regression models after nearest-neighborhood matching, maintaining a case (IC with CAD) and treated-control (CAD without IC) ratio 1:3. ^30^
b. Age ≥ 75 yrs: To further account for age, we restricted our analysis to participants with age ≥ 75 yrs.^31^ c)
c. AoU v8 latest data release (C2024Q3R9, Feb 2026) analysis: To further account for lipid levels and systolic blood pressure (SBP), we identified CAD participants (CAD=22,089, including IC=1,343) with consent age ≥ 60 years and APO (ε) genotyping using AoU v8 latest data release (C2024Q3R9, Feb 2026).

We further excluded CAD participants without documented lipid levels and SBP at enrollment based on the corresponding concept ID (Details in Supplement 1). Finally, we obtained 17,050 CAD participants (including IC=1,141). For each participant, we obtained baseline lipid levels (total cholesterol (TC), low-density lipoprotein (LDL), and high-density lipoprotein (HDL)) and the latest lipid levels, and calculated delta TC, delta LDL, and delta HDL (the difference between baseline and latest lipid levels). Among CAD participants with lipid levels measured (n=17,050), 75% had baseline lipids assessed before the onset of CAD, and 95% had the latest lipids after the onset of CAD. Among IC (n=1,141), 98% had baseline lipids before the onset of IC, and 70% had the latest lipids after the onset of IC. We recorded the lowest systolic blood pressure at enrollment. For the multivariable model, we included additional confounding comorbidities, such as obstructive sleep apnea, chronic kidney disease, and community deprivation index, a composite score of six census tract-level variables from the 2017 American Community Survey (Details in Supplement 1). We subsequently assessed the association of CABG with IC after adjusting for clinical factors (age, sex, race/ethnicity, diabetes, hypertension, hyperlipidemia, depression, ischemic stroke, obesity (BMI ≥ 30), the smoking, alcohol use, obstructive sleep apnea, chronic kidney disease, statin use, antihypertensive use, antidiabetic use), social determinants (health insurance, education (college graduate and/or advanced degree), annual household income, employment status, community deprivation index), delta lipids, SBP, and APO (ε) genotypes.

#### 3) Statistical tools

We performed all analyses in R, integrated within Jupyter Notebooks (a secure, access-controlled, cloud-based analysis environment) on the AoU researcher workbench. We used the matchit and samplesizeestimator packages for propensity-score matching and sample size estimation.^30^ For primary analyses, we estimated 3% probability of IC in the non-exposed group, and to detect a minimum odds ratio (RR) of 1.30 for CABG (exposed), we calculated ∼2600 CABG cases (1:1 exposed: unexposed ratio) to be required for achieving an alpha level of 5%. We defined the nominal statistical significance of p-values for all analyses at 2-sided p-values <□ 0.05.

## RESULTS

We screened 34,161 participants with age ≥ 60 years and with at least one hospital or outpatient encounter, with ICD9/10 codes corresponding to myocardial infarction, stable or unstable angina, chronic ischemic heart disease, percutaneous coronary intervention, or CABG (**Figure 1**). We further excluded 2,054 participants (related individuals (n=2024); call rate <95%(n=30)) after genotype quality control filtering, 9,437 participants with fewer than 2 visit encounters corresponding to CAD, and 321 participants with prevalent IC. Finally, we included 22,349 CAD participants (IC with CAD = 908; CAD without IC = 21,441) in our final analysis. **Supplement 1** represents enrollment counts from 2017 to 2023.

### 1) Baseline characteristics

When we compared baseline characteristics (**Table 1**), we noted that participants with IC and CAD were significantly older (77.83 years vs 73.8 years, p<0.001) than those with CAD alone. When we compared racial and ethnic groups, we observed higher proportions of self-reported Black race (15.0% vs 11.9%, p=0.007) and Hispanic ethnicity (11.3% vs 8.8%, p=0.008) in IC participants than in CAD-only participants. CAD participants had a mean follow-up of 8 years (IQR 4-11 years) till 2023, and the mean difference between the onset of IC and CAD was 7.3 years (IQR 3-10 years). IC had a significantly higher proportion of clinical factors than CAD. IC had higher proportions of APO ε3ε4 (29.5% vs 21.6%, p < 0.001) and APO ε4ε4 genotypes (3.9% vs 2.0%, p < 0.001) than CAD. Among subtypes of IC, we observed the highest proportion in mild cognitive impairment (54%), followed by other types of dementia (26%), vascular dementia (10%), and Alzheimer’s dementia (AD) (5%) (**Table 1**)

**Table 1:** Baseline characteristics (AllofUS, v8, Age ≥60)

|  | CAD without IC (n=21,441) | IC with CAD (n=908) | P value |
| --- | --- | --- | --- |
| **Demographics\*** |  |  |  |
| Age (mean, SD) | 73.85 (60-104, SD=7.9) | 77.83 (60-100, SD=8.3) | **p<0.001** |
| Female sex | 8583 (40%) | 366 (40.3%) | p=0.867 |
| Health insurance | 20,739 (96.7%) | 870 (95.8%) | p=0.133 |
| **Race/ethnicity** |  |  |  |
| White race | 15,032 (70.1%) | 583 (64.2%) | **p<0.001** |
| Black race | 2,561 (11.9%) | 136 (15.0%) | **p=0.007** |
| Other | 1,968 (9.2%) | 86 (9.5%) | p=0.81 |
| Hispanic | 1880 (9.2%) | 103 (11.3%) | **p=0.008** |
| **Household income** |  |  |  |
| Not reported | 4630 (21.6%) | 250 (27.5%) | **p<0.001** |
| Less than equal to \$25,000 | 4169 (19.4%) | 218 (24.0%) | **p<0.001** |
| Between \$25,000-\$75,000 | 5954 (27.8%) | 247 (27.2%) | p=0.73 |
| More than \$75,000 | 6688 (31.2%) | 193 (21.3%) | **p<0.001** |
| **Employment** |  |  |  |
| Not reported | 340 (1.6%) | 23 (2.5%) | **p=0.04** |
| Unemployed | 3717 (17.3%) | 142 (15.6%) | p=0.20 |
| Retired | 12,237 (57.1%) | 624 (68.7%) | **p<0.001** |
| Employed | 5,147 (24.0%) | 119 (13.1%) | **p<0.001** |
| **Education** |  |  |  |
| Highest Education | 10,009 (46.7%) | 377 (41.5%) | **p=0.002** |
| **Clinical factors** |  |  |  |
| Depression | 8,102 (37.8%) | 588 (64.8%) | **p<0.001** |
| Ischemic Stroke | 2,491 (11.6%) | 265 (29.2%) | **p<0.001** |
| Hyperlipidemia | 19,805 (92.4%) | 883 (97.2%) | **p<0.001** |
| Hypertension | 19,379 (90.4%) | 870 (95.8%) | **p<0.001** |
| Diabetes | 10,057 (46.9%) | 518 (57.0%) | **p<0.001** |
| Obesity | 9,729 (45.4%) | 388 (42.7%) | p=0.117 |
| Smoking | 5,299 (24.7%) | 245 (27.0%) | p=0.121 |
| Alcohol Use | 1,992 (9.3%) | 115 (12.7%) | **p<0.001** |
| Statin use | 16,438 (76.7%) | 795 (87.6%) | **p<0.001** |
| Antihypertensive use | 18,503 (86.3%) | 830 (91.4%) | **p<0.001** |
| Antidiabetic use | 10,161 (47.4%) | 515 (56.7%) | **p<0.001** |
| CABG | 7721 (36%) | 414 (45.6%) | **p<0.001** |
| **APO (ε) genotypes** |  |  |  |
| **APOε2ε2** | 132 (0.6%) | 5 (0.6%) | p=0.97 |
| **APOε2ε3** | 2,478 (11.6%) | 90 (9.9%) | p=0.14 |
| **APOε3ε3** | 13,312 (62.1%) | 484 (53.3%) | **p<0.001** |
| **APOε2ε4** | 468 (2.2%) | 26 (2.9%) | p=0.21 |
| **APOε3ε4** | 4,629 (21.6%) | 268 (29.5%) | **p<0.001** |
| **APOε4ε4** | 422 (2.0%) | 35 (3.9%) | **p<0.001** |
| **IC type** |  |  |  |
| **Mild cognitive impairment** | NA | 540 (59.5%) |  |
| **Alzheimer's dementia** | NA | 42 (4.6%) |  |
| **Vascular dementia** | NA | 92 (10.1%) |  |
| **Other dementia** | NA | 234 (25.7%) |  |

\*Proportions in bold with P value <0.05 (Pearson's Chi-squared test, baseline group = CAD)
Obesity (Body mass index ≥ 30). CAD: Coronary Artery Disease; IC: Impaired Cognitive Disease; CABG: Coronary Artery Bypass Graft
Highest Education: College graduate and/or advanced degree

**Table 2:** Proportions of IC in CAD based on APO (ε) genotype, AllofUS v8, Age ≥60.

|  | No CABG<br>(n=14,214) | CABG<br>(n=8,135) | P Value* |
| --- | --- | --- | --- |
| APO (ε) variants |  |  |  |
| APOε2ε2 | 5/91 (5%) | 0/46 (0%) | NA |
| APOε2ε3 | 41/1693 (2.4%) | 49/875 (5.6%) | **p=5.4e-05** |
| APOε3ε3 | 275/8771 (3.1%) | 209/5025 (4.1%) | **p=0.001** |
| APOε2ε4 | 13/320 (4.0%) | 13/174 (7.5%) | p=0.15 |
| APOε3ε4 | 136/3047 (4.4%) | 132/1850 (7.1%) | **p=8.8e-05** |
| APOε4ε4 | 24/292 (8.2%) | 11/165 (6.6%) | p=0.67 |

\* Bold values with higher proportions having p value ≤0.05
CAD: Coronary Artery Disease;
IC: Impaired cognitive disease defined as all-cause dementia or mild cognitive impairment
CABG: Coronary artery bypass graft

### 2) Association of CABG with IC

Among all CAD participants, we observed that the proportion of IC was significantly higher in CABG participants (**5.1% vs 3.5%, p=1e-08**) than in non-CABG participants (**eTable 2**). In the multivariable logistic regression model after adjusting for demographics, clinical factors, social determinants, and APO (ε) genotype groups, CABG remained significantly associated with IC (**1.23;1.06-1.41, p = 0.005**). Among clinical factors, both depression (**3.13;2.7-3.64, p=2e-16**) and ischemic stroke (**2.31;1.97-2.70, p=2e-16**) were strongly associated with IC in CAD participants. Among APO (ε) genotype groups, we observed a strong association of APO ε4ε4 (**2.54;1.71-3.63, p=1.0e-06**), APO ε3ε4 (1.**62;1.38-1.90, p =2.3e-09**) with IC in CAD participants. However, when we stratified CAD participants based on APO (ε) genotype groups, we observed the highest association of CABG with IC in the ε2/ε3 group (**1.91;1.21-3.02, p = 0.005**). (Table 3) Our association results did not change when we used genetically predicted ancestries (data not shown). 3) EtM survey data:

**Table 3:**
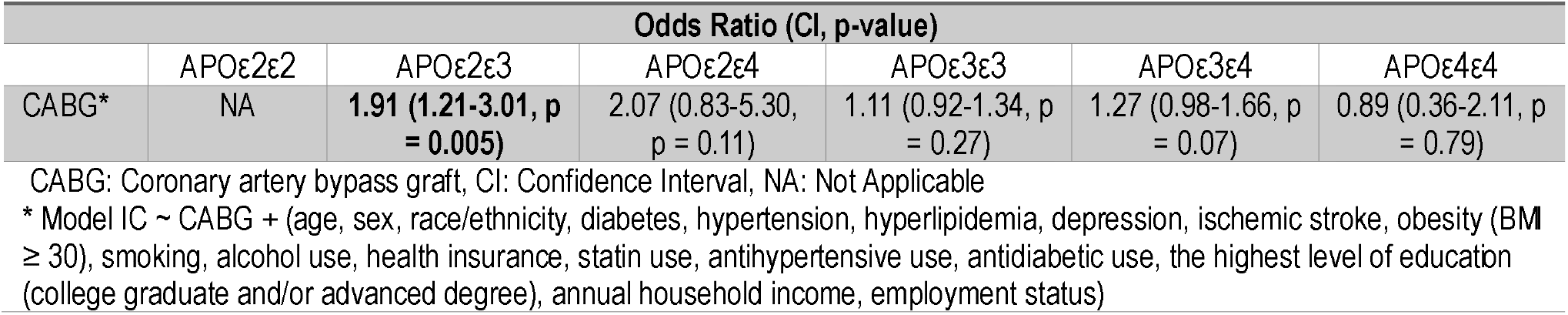
Association of CABG with IC based on APO (ε) genotypes in CAD, AllofUS v8, Age ≥60.

| Odds Ratio (CI, p-value) |  |  |  |  |  |  |
| --- | --- | --- | --- | --- | --- | --- |
|  | APOε2ε2 | APOε2ε3 | APOε2ε4 | APOε3ε3 | APOε3ε4 | APOε4ε4 |
| CABG* | NA | **1.91 (1.21-3.01, p = 0.005)** | 2.07 (0.83-5.30, p = 0.11) | 1.11 (0.92-1.34, p = 0.27) | 1.27 (0.98-1.66, p = 0.07) | 0.89 (0.36-2.11, p = 0.79) |

CABG: Coronary artery bypass graft, CI: Confidence Interval, NA: Not Applicable
\* Model IC ~ CABG + (age, sex, race/ethnicity, diabetes, hypertension, hyperlipidemia, depression, ischemic stroke, obesity (BMI ≥ 30), smoking, alcohol use, health insurance, statin use, antihypertensive use, antidiabetic use, the highest level of education (college graduate and/or advanced degree), annual household income, employment status)

When we compared GradCPT (d-prime) scores in CAD participants with and without CABG, we observed lower scores in those with CABG (**Table 4**). When we performed linear regression to assess the association of CABG with GradCPT scores after adjusting for age, race/ethnicity, sex, APO (ε) genotype, stroke, depression, smoking, alcohol use, health insurance, income, employment and educational status, we observed a consistent negative association of CABG with GradCPT score (**beta estimate = -0.09, p=0.04**).

**Table 4:** Exploring the Mind, AllofUS, v8, Age ≥60^1^.

| Gradual Onset Continuous Performance Task: GradCPT (City or Mountain) |  |  |  |
| --- | --- | --- | --- |
|  | No CABG (n=696) | CABG (n=302) | P Value* |
| GradCPT: d-prime, Mean (SD, range) | 2.33 (0.76, 0.56-4.76) | **2.18 (0.72, 0.56-4.23)** | **p=0.004** |

\*Bold values with higher proportions having p value <0.01; CABG: Coronary artery bypass graft
<sup>1</sup>Includes all coronary artery disease participants who completed the optional GradCPT test in 2023
GradCPT assesses core executive function by testing sustained attention and inhibitory control.
d-prime: a measure of discrimination ability that represents the participant's ability to withhold responses to mountains while making responses to the more prevalent city images

### C) Sensitivity Analysis

#### 1) Propensity Score Matched Analyses

**e Table 2 in Supplement 1** describes baseline characteristics of IC and CAD after propensity matching, and **eFigure 2** in **Supplement 1** reports the corresponding standardized mean difference propensity matched plots. The SMD of various matching variables between IC (n=908) and CAD (n=21,441) was 0.64 which reduced to 0.04 after matching between IC (n=908) and CAD (2,724). After propensity-score matching for demographics, clinical factors, social determinants, and APO (ε) genotypes between IC and CAD, we observed a persistent association of CABG with IC (**1.21;1.04-1.41, p =0.01**)

#### 2) Age ≥ 75 yrs

The association of CABG with IC persisted when we restricted to participants with age ≥ 75 yrs (**1.32;1.10-1.57, p =0.002**) after adjusting for demographics, clinical factors, social determinants, and APO (ε) genotypes.

#### 3) AoU v8 latest data release (C2024Q3R9, Feb 2026) analysis

Among CAD participants with lipid levels (n=17,050) the mean age of participants when their baseline lipid levels were recorded was lower than their latest lipid levels (62 ± 9.6 yrs vs 73 ± 7.6 yrs, p<0.001). Among all CAD participants, mean TC (deltalTC (decrease): 31 ± 48 mg/dl) and mean LDL levels (deltaLDL (decrease): 27 ± 41 mg/dl) decreased, and mean HDL levels increased (deltaHDL(increase): (1 ± 11) mg/dl), respectively, from baseline to the latest lipid levels. The association of CABG with IC persisted (**1.24;1.10-1.42, p=0.0009**) in the AoU latest data release after adjusting for clinical factors, social determinants, APO (ε) genotype, delta lipids, and SBP (Table 5).

**Table 5:** Association of CABG with IC in CAD, AllofUS v8, Age ≥60.

| Odds Ratio (CI, p-value) |  |  |
| --- | --- | --- |
|  | AoU data release (C2024Q3R5, released Feb 2025) | AoU data release (C2024Q3R9, Feb 2026) |
| CABG | 1.23(1.06-1.41, p = 0.005)* | 1.24 (1.10-1.42, p=0.0009)† |

CABG: Coronary artery bypass graft, CI: Confidence Interval, NA: Not Applicable
\* Model IC ~ CABG + APO (ε) genotypes + (age, sex, race/ethnicity, diabetes, hypertension, hyperlipidemia, depression, ischemic stroke, obesity (BMI ≥ 30), smoking, alcohol use, health insurance, statin use, antihypertensive use, antidiabetic use, the highest level of education (college graduate and/or advanced degree), annual household income, and employment status)
† Model IC ~ CABG + APO (ε) genotypes + delta lipids (difference between baseline and latest total cholesterol, low density lipoprotein, and high density lipoprotein) + lowest systolic blood pressure (at the time of enrollment) + (age, sex, race/ethnicity, diabetes, hypertension, hyperlipidemia, depression, ischemic stroke, obesity (BMI ≥ 30), obstructive sleep apnea, chronic kidney disease, smoking, alcohol use, health insurance, statin use, antihypertensive use, antidiabetic use, the highest level of education (college graduate and/or advanced degree), annual household income, employment status, and community deprivation index)

## DISCUSSION

In our large and diverse retrospective study of AoU CAD participants (age ≥ 60 yrs), we observed an association between CABG and IC, i.e., mild cognitive impairment and/or all-cause dementia, after adjusting for demographics, clinical factors, social determinants, and APO (ε) genotype. We further noted that the strongest association between CABG and IC was observed in the APO ε2/ε3 genotype group. In AoU participants who had completed the EtM survey, we noted that CABG was associated with lower neurocognitive GradCPT scores. To our knowledge, this is the first study to investigate the association of CABG with IC and neurocognitive test scores across APO (ε) genotype groups in a diverse participant group.

The association between CABG and IC has been controversial, with mixed results.^3,6,9,10,32,33^ Many CABG studies evaluating cognitive decline suffer from a lack of power and a lack sex/racial diversity.^9,10^ We tried to define IC to conform with the POCD definition by Evered et al by including an age cut off, having either mild cognitive impairment (MCI) or dementia, and requiring at least two or more health care visit encounters related to IC.^27^ Evered et al. highlight many similarities between MCI and POCD.^27^ While MCI has been reported to have a bidirectional cognitive evolution trajectory, 14%-18% per year evolve into dementia, and the increase is likely exacerbated by underlying vascular risk factors.^27,34,35^ Our large sample size (CABG, n∼8000) and diverse dataset overcome some of these challenges. Consistent with prior CAD cohorts, we observed lower proportions of Alzheimer’s dementia in our CAD group, however, we may be under-reporting the IC proportions in general as our EHR follow up ended in 2023.^4^ However, we observed persistent strong association APO ε3/ε4, ε4ε4 genotypes with IC across all CAD participants suggesting shared burden of APO ε4 polymorphism related neurodegenerative mechanisms and vascular risk factors in IC participants having CAD.^16,17,36,37^ A recent study evaluating the association of APO(ε) genotypes with Alzheimer’s dementia noted co-existing vascular brain injury in more than 80% of cases.^37^ While we observed an independent association of CABG with IC in CAD participants after adjusting for APO (ε) genotypes, we noted the highest association in the APO ε2/ε3 group. Our observations suggest non-APO ε4 mechanisms in CABG-associated IC and are consistent with other CABG studies evaluating the association of APO ε4 with postoperative cognitive decline.^38,39^ The association of APO ε4 genotype with amyloid beta plaque burden in AD relative to APO ε2 genotype has been extensively investigated.^16,37,40^ However, a recent study of AD patients by Reiman et al. reported 52% (75/143) with APO ε2/ε3 genotype had co-existing vascular brain injury compared to 45% (482/1065) with APO ε3/ε3 genotype, and 35% (419/1174) with APO ε3/ε4 genotype.^37^ The authors also reported a trend towards association of APO ε2/ε3 with AD patients having vascular brain injury (unadjusted odds=1.33;0.94 - 1.89, p=0.11).^37^ Further, Zhao et al reported APO ε2 mice had increased tau phosphorylation compared to APO ε4 mice when injected with human tau protein in the lateral ventricle using adeno-associated virus.^41^ Finally, more recently, Ojo et al assessed inferior frontal gyrus cerebrovascular proteomic expression of AD cases and healthy controls across different APO (ε) genotypes.^42^ In that study, while AD patients with APO (ε2) genotypes had lower amyloid beta plaque burden and neurofibrillary tangle score than APO (ε4) genotype AD patients, APO (ε2) genotype AD patients had a distinct cerebrovascular transcriptional and proteomic profile compared with APO (ε4) genotype AD patients. APO (ε2/ε3) genotypes showed upregulation of DNA methylation, the spliceosomal cycle, transcriptional repression signaling, and the Base excision repair (BER) pathway. In contrast, APO ε3/ε4 and ε4ε4 genotypes showed upregulation of EIF2, eIF4, and p706k signaling, as well as the mTOR pathway, in the cerebral vasculature.^42^ These observations suggest alternative neurodegenerative mechanisms in the APO ε2/ε3 genotype in CABG-associated IC.^5,33^ Further, APO (ε) genotype frequencies differ across ancestries, exhibit heterogeneity in phenotypic associations based on ancestries, and APO (ε) genotype-defining SNVs (rs429359 and rs7412) exhibit differential colocalization with other IC associate SNVs.^12,13^ Hence, future studies evaluating APO (ε) genotype interaction with ancestries in CAD can provide pathophysiologic insight and identity risk reduction strategies across ancestries.

We observed that the association between CABG and IC persisted after propensity matching, particularly among participants aged≥ 75 years. Further, our EtM data showed persistent low GradCPT scores in the CABG group (**Table 4**) even after adjusting for demographics, social determinants, and APO (ε) genotype. Unlike many studies evaluating short-term postoperative cognitive scores in CABG patients, in our study, the CABG participants who completed the EtM survey had a mean time difference between onset of CABG and survey date of 5.9 years (SD=5.3).^9^ In this regard, our study provides insight into medium to long term cognitive scores after CABG. Further, we included GradCPT as our secondary outcomes to support the POCD construct, as neurocognitive testing to assess executive function has been included in the POCD definition.^27^

Finally, considering our observations, our study raises case for genetic evaluation in CAD patients for IC risk assessment. Currently, comprehensive IC and CAD combined genetic risk panels that include APO (ε) genotyping are readily and commercially available at affordable costs, which can be used either at the time of diagnosis of CAD or counseling prior to CABG or used as screening tools post CABG.^43^ Future basic mechanistic studies linking the APO (ε) genetic architecture in CABG and IC can help us identify risk-reduction strategies for the CABG subgroup.

### Limitations

Our retrospective study design is subject to limitations, such as information and measurement bias. We defined IC using ICD/SNOMEDS codes for dementia and mild cognitive impairment to capture the entire spectrum of cognitive impairment. Given that enrollment in AoU required informed consent, participants with severe IC phenotypes at the time of consent were likely not included. We acknowledge that our IC definition exhibits heterogeneity; however, IC clinical phenotypes often overlap and share heterogeneous neuropathologies and frequently have a high burden of vascular brain injury.^37^ Further, we increased the likelihood of an accurate diagnosis by restricting to patients aged ≥ 60 years, performing a sensitivity analysis in patients aged ≥75 years, and including EtM survey data.^20,31^ Similarly, we aimed to capture the entire spectrum of CAD, and our CAD definition has some heterogeneity. However, our definition is consistent with previously published CAD studies.^11,23^ We used an EHR-based index date of diagnosis for our phenotypes and acknowledge that there could be some discordance between the “true onset of disease” and the EHR-recorded diagnosis date.^44^ We also acknowledge that both CAD and IC are complex phenotypes with significant preclinical evolution that may not be captured adequately in EHR based studies.^10,13,29^ Given AoU typically integrates EHR data and not claims data, the accuracy of chronic diagnoses dates remains higher than procedural dates, hence there could be some discordance between CABG and IC onset date. Considering these limitations, we only noted 8% of IC cases preceded the recorded CABG dates.^45^ While we had adequate power (>80%) for our primary analyses, we did not have enough power for all subgroup analyses. We also acknowledge the possibility of confounding by indication and CAD severity when evaluating the association of CABG with IC, and unmeasured confounders can affect our association analyses. However, we sought to limit their impact by using propensity score matching of multiple confounders, including social determinants, and further adjusting for APO (ε) genotype. We further confirmed the association of CABG with IC using the AoU latest release (C2024Q3R9, Feb 2026) by adjusting for additional confounding variables (Table 5). Moreover, our study design, using the AoU dataset and a standardized data collection model that integrates decades of EHR data for each patient, can somewhat mitigate these biases and serve as a stepping stone for future prospective studies in this field.

In conclusion, we identified CABG to be significantly associated with IC after CAD, after adjusting for APO (ε) genotype. When stratified by APO (ε) genotype status, we observed the highest association of CABG with IC in the APO ε2/ε3 group. Our observations highlight the role of APO (ε) genotype evaluation in CAD patients for IC risk assessment and provide a path for future mechanistic studies evaluating the association of IC with CABG.

## Data Availability

All data produced are available online at

## Acknowledgments

We gratefully acknowledge AllofUs participants for their contributions, without whom this research would not have been possible. We also thank the NIH AllofUs Research Program for making available the participant data examined in this study.

## Author Contributions

The team had full access to all of the data in the study and take responsibility for the integrity of the data and the accuracy of the data analysis.

## Concept and Design: All authors

Acquisition, analysis, or interpretation of data: Dr.Hariharan, Dr. Asamoah, and Dr.Bagheri Drafting of the initial manuscript: Dr.Hariharan

Statistical analysis: Dr.Hariharan, and Dr.Bagheri

Critical review of the manuscript for important intellectual content: Dr.Pottinger, Dr.Machipisa. Dr.Singh, Dr.Voiculescu, Prof A Opekun, Dr.Josee Dupuis, Dr.J Dawn Abbott and Dr.Frank Sellke.

Disclaimer: The views expressed are those of the authors and not necessarily those of the funders, NIH, or AllofUS research program.

## Consent Statement

All patients were enrolled in the AoU research program after informed consent.

## Ethics Statement

Patient consent for publication: Not applicable.

Ethics approval: Brown University Research Agreements and Contracting Committee reviewed our retrospective analysis proposal and approved the use of AoU researcher workbench. In addition, we have reviewed, completed AoU data access requirements and are in compliance with the data user code of conduct for registered and controlled tier data. There is no human/animal involved in this retrospective record review study and hence it does not need to be reviewed and approved by the AoU IRB.

