## Supplement 1 for "Association of coronary artery bypass with cognitive impairment in coronary artery disease across APO (ε) genotypes in AllofUS"

Supplement 1 (Index)

1) Case definitions

2) Clinical factors and social determinants definitions.

3) eFigure 1: AoU enrollements from 2017-2023

4) eTable1: Proportions of IC in AllofUS, v8 with CAD stratified by CABG, Age ≥60

5) eFigure 2: Propensity matched plot of IC and CAD

6) eTable 2: Baseline characteristics in IC vs CAD (AllofUS, v8, Age ≥60) before and after propensity matching

Case definitions

(OMOP Standard Concept Names)

A) Impaired Cognitive disease (IC) had following electronic health records (EHR) diagnoses:

**ICD 9 codes:**

Dementia or MCI: ‘290.0', '290.10', '290.11', '290.12', '290.13', '290.20', '290.21', '290.3', '290.40', '290.41', '290.42', '290.43', ‘290.8’, ‘290.9’, ‘291.2’, ‘294.0’,'294.1', '294.10', '294.11', ‘294.20’, ‘294.21’,'331.0', ‘331.7’,'331.11', '331.19',’331.2’,’331.83’

**ICD 10 codes:**

Dementia or MCI: 'F01.50', 'F01.51', ‘F01.52’, ‘F01.A0’, ‘F01.B0’, ‘F01.C4’,‘F02.80’, ‘F02.81’,’F02.811’, ‘F02.818’, ‘F02.A0’, ‘F02.A18’, ‘F02.B0’, ‘F02.B3’, ‘F02.B4’, ‘F03.90', 'F03.91', ‘F03.911’, ‘F03.918’, ‘F03.92’, ‘F03.92’, ‘F03.93’, ‘F03.94’, ‘F03.A0’, ‘F03.A18’, ‘F03.B18’, ‘F03.C3’, ‘G30.1’, ‘G30.8’, 'G30.9', 'G31.01', 'G31.84', ‘G31.84’, 'F10.27'

1. Alzheimer's disease
2. Amnestic disorder
3. Dementia
4. Dementia associated with alcoholism
5. Dementia associated with another disease
6. Dementia with behavioral disturbance
7. Frontotemporal dementia
8. Mild cognitive disorder
9. Mild dementia
10. Minimal cognitive impairment
11. Moderate dementia
12. Multi-infarct dementia with depression
13. Multi-infarct dementia, uncomplicated
14. Presenile dementia with depression
15. Primary degenerative dementia of the Alzheimer type, presenile onset
16. Primary degenerative dementia of the Alzheimer type, senile onset
17. Senile degeneration of brain
18. Senile dementia with depression
19. Uncomplicated presenile dementia
20. Uncomplicated senile dementia
21. Vascular dementia with behavioral disturbance
22. Vascular dementia without behavioral disturbance

B) Coronary Artery Disease (CAD) had following electronic health records (EHR) diagnoses:

**ICD 9 codes:** ‘413.0’, ‘413.1’, ‘413.9’, ‘414’,’414.0’, ‘414.00’, ‘414.01’, ‘414.02’, ‘414.03’, ‘414.04’

‘414.05’, ‘414.07’, ‘414.10’, ‘414.11’, ‘414.12’, ‘414.19’, ‘414.2’, ‘414.3’, ‘414.4’, ‘414.8’, ‘414.9’

**ICD 10 codes:** ‘I20’, ‘I20.0’, ‘I20.1’, ‘I20.8’, ‘I20.9’, ‘I21’, ‘I21.01’, ‘I21.02’, ‘I21.09’

‘I22.1’, ‘I22.2’, ‘I23.1’, ‘I23.3’, ‘I23.5’, ‘I23.6’, ‘I23.8’, ‘I24.0’, ‘I24.1’, ‘I24.8’,

‘I24.9’, ‘I25’, ‘I25.1’, ‘I25.10’, ‘I25.11’, ‘I25.110’, ‘I25.111’, ‘I25.118’ , ‘I25.119’, ‘I25.2’

‘I25.3’, ‘I25.41’, ‘I25.42’, ‘I25.5’, ‘I25.6’, ‘I25.700’, ‘I25.701’, ‘I25.708’, ‘I25.709’, ‘I25.710’,

‘I25.718’, ‘I25.719’, ‘I25.720’, ‘I25.721’, ‘I25.729’, ‘I25.730’, ‘I25.738’, ‘I25.758’, ‘I25.759’, ‘I25.760’

‘I25.768, ‘I25.790’, ‘I25.810’, ‘I25.812’, ‘I25.82’, ‘I25.83’, ‘I25.84’, ‘I25.89’, ‘I25.9’

1) Chronic total occlusions of coronary artery
2) Acute ischemic heart disease
3) Chronic ischemic heart disease
4) Coronary artery thrombosis
5) Coronary arteriosclerosis
6) Acute coronary artery occlusion not resulting in myocardial infarction
7) Coronary artery atherosclerosis
8) Coronary artery bypass graft finding
9) Coronary thrombosis not resulting in myocardial infarction
10) Myocardial infarction due to demand ischemia
11) Angina pectoris
12) Acute ST elevation myocardial infarction
13) Coronary artery bypass graft occlusion
14) Atherosclerosis of coronary artery without angina

Definitions of predictors and co-variates

(OMOP Standard Concept Names with SNOMED Concept ID)

A) Coronary artery bypass graft (CABG):

a) Aortocoronary bypass graft present (42537729)

- Coronary artery bypass graft finding (312922)
- Coronary artery bypass graft present (42537730)

b) Arteriosclerosis of coronary artery bypass graft (443563)

B) Clinical factors:

1) Body Mass Index (BMI):

- Earliest BMI value obtained from electronic medical records (3038553)

2) Depression: (440383, 4152280)

- Acute depression
- Atypical depressive disorder
- Bipolar affective disorder, current episode depression
- Chronic depression
- Chronic depressive personality disorder
- Depressive disorder in remission
- Dysthymia
- Major depressive disorder
- Mild, moderate, severe and recurrent depression
- Schizoaffective disorder, depressive type

3) Ischemic stroke: (4111710, 443454, 381316, 4153352, 4310996, 4046360)

- Brainstem stroke syndrome
- Cerebral infarction
- Cerebrovascular accident
- Embolic stroke
- Ischemic stroke
- Lacunar infarction

4) Hypertension: (312648, 4028741, 4167358, 320128, 317898, 4209293)

- Benign hypertension
- Essential hypertension
- Malignant essential hypertension
- Diastolic and Systolic hypertension

5) Hyperlipidemia: (44834563, 432867, 35207065, 438720, 35207062, 35207061)

- Endogenous hyperlipidemia
- Hypercholesterolemia
- Hypertriglyceridemia
- Mixed hyperlipidemia
- Secondary hyperlipidemia

6) Diabetes: (201820, 44833365, 1567940, 201826, 1567956)

- Diabetes during pregnancy
- Diabetes without complication
- Secondary diabetes
- Type 1 and 2 diabetes

9) Alcohol use: (4218106)

- Alcohol abuse
- Alcohol dependence
- Chronic alcoholism in remission
- Chronic continuous alcoholism
- Episodic chronic alcoholism

10) Smoking: (4209423)

- Nicotine dependence
- Nicotine dependence in remission
- Tobacco dependence syndrome

11) Obstructive sleep apnea: (442588)

- Mixed sleep apnea
- Obstructive sleep apnea of adult

12) Chronic kidney disease: (46271022, 443614, 443601, 443597, 45763854, 443611, 443612, 4030520)

- Chronic kidney disease stage 1-5
- End-stage renal failure on dialysis.

13) Statin drugs: (21601855)

14) Antihypertensive drugs: (21600381)

- Betablockers (21601664)
- Renin-angiotensin system inhibitors (21601782)
- Diuretics (21601461)
- Peripheral vasodilators (21601560)
- Calcium channel blockers (21601744)

15) Antidiabetic drugs

- Fast acting insulin (51428, 400008, 86009, 314686, 253182, 221109)
- Intermediate acting insulin (314684)
- Long acting insulin (1670007, 139825, 274783)
- Insulin analogues for inhalation (631657)
- Biguanides (6809, 8129)
- Sulfonylurea (2404, 25789, 4821, 4815, 10633, 10635)
- Alphaglucosidase inhibitors (16681, 30009)
- Thiazolidiendiones (33738, 84108, 72610)
- Dipeptidyl peptidase 4 inhibitors (1368001, 1100699, 857974, 593411)
- Glucagon like peptide-1 inhibitors (1534763, 1551291, 60548, 475968, 1440051, 1991302)
- Sodium-glucose- cotransport 2 inhibitors (1373458, 1488564, 1545653, 1992672)

C) Social determinants:

1) Annual household income: (1585375)

- Baseline reference annual income (>USD $75,000)

2) Employment: (1585952)

- Baseline reference employment status (unemployed or unable to work)

3) Health insurance status (585386):

- No/Skip/Prefer not to answer (Group = 0)
- Yes (Group = 1)
- Don’t know (Group = 2)

4) Community Deprivation index (composite score based on the following census tract variables):

- Poverty fraction: Fraction of households with income below poverty level within the past 12 months
- Median income: Median household income in the past 12 months in 2017 inflation-adjusted dollars
- Fraction high school: Fraction of population 25 and older with educational attainment of at least high school graduation (includes GED equivalency)
- Fraction insured: fraction of population with health insurance
- Fraction SNAP: Fraction of households receiving public assistance income or food stamps/SNAP in the past 12 months
- Fraction vacant: Fraction of houses that are vacant

D) Lipid level data:

1) Total Cholesterol (TC):

Concept ID: 4260765, 44791053, 4190897, 40484105, 4008265,3027114,40757569,2212267,4260765, 44791053, 4190897, 40484105, 4008265, 3037598, 40757569, 4260765, 44791053, 4190897, 3002651, 2212267,44787078, 40484105, 4008265, 40780291,3027114, 40757569,2212267, 3024723,3033364, 3024723, 3033364, 3015232

2) Low density lipoprotein (LDL):

Concept ID: 3008631,3028437,3009966, 3035899, 3028288,3053341, 2212450, 3049222, 3050458, 40760809, 3035009, 3049237, 3052982, 3028288, 3046493,3001308, 3045323, 3009966, 4191837, 4041721, 4042062, 40795800, 3033200, 42870529, 3050730, 3030437, 3038988, 3008631, 3035899, 4210878, 4041556, 4042061, 4042758

3) Hight density lipoprotein (HDL):

Concept ID: 3007070,3050630,3032633,3011163, 3009718, 4041557, 3053286, 4041720, 4042059, 4042081, 4055665, 4076704, 4101713, 4195503, 3003767, 3005561, 3033011, 3007070, 3009718,3 011884, 3013473, 3015204, 3020107, 3020189, 3022449,3023574,3023602,3023752,3024401,3030792, 40761042, 40757503, 40782589,3032771, 3033190, 3033638, 3034482,3040324,3040815, 40759254, 3050988,40798534, 40795258, 40795251, 40785417, 40789378, 40795256, 40782086, 40788729, 3033190, 3020189, 3023602, 3009718, 40761043, 40759253, 42868674, 40758736, 3047111,3050038, 40757599, 4021289, 40778721, 40775398, 40795257, 40795254, 40788728, 40795253, 40785416, 40782761, 40778719

C) Systolic Blood Pressure (SBP):

Concept ID: 4232915, 4292062, 4161413, 4248525, 4197167, 37394652, 37396683, 3012311, 3025407, 3033616, 3025048, 3025957, 3011870, 3035856, 21492239, 3018586, 903114, 903118, 46284421, 4152194, 3013880, 3009395, 3011579, 3010693, 3006833, 3013443, 903130, 903109,3004249

eFigure 1: AllofUS enrollements per year

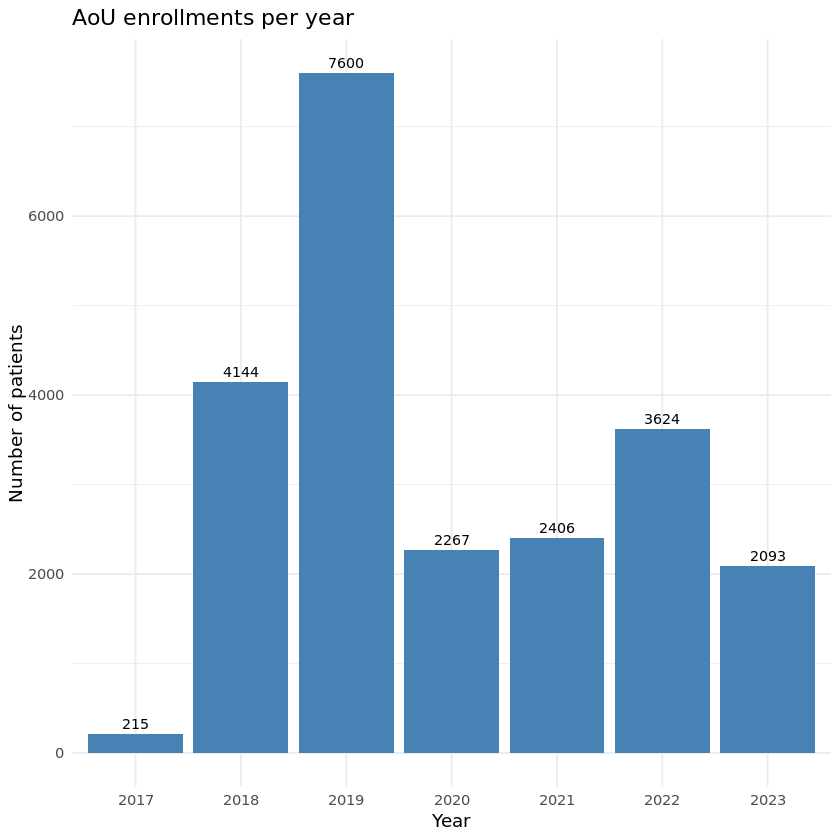

| e Table 1: Proportions of IC in AllofUS, v8 with CAD stratified by CABG, Age ≥60 | | | |
| --- | --- | --- | --- |
|  | No CABG  (n=14,214) | CABG  (n=8,135) | P Value* |
| IC | 499 (3.5%) | **414 (5.1%)** | **p=1e-08** |
| IC type | | | |
| a) Mild Cognitive Impairment | 290 (2.0%) | **250 (3.1%)** | **p=1e-06** |
| b) Alzheimer’s Dementia | 23 (0.1%) | 19 (0.2%) | p=0.30 |
| c) Vascular Dementia | 50 (0.3%) | 42 (0.5%) | p=0.08 |
| d) Other Dementia | 136 (0.9%) | **103 (1.3%)** | **p=0.03** |
| * Bold values with higher proportions having p value ≤0.05  CAD: Coronary Artery Disease;  IC: Impaired Cognitive Disease defined as all-cause dementia or mild cognitive impairment. All events captured via electronic health records till 2023.  CABG: Coronary artery bypass graft | | | |

eFigure 2: Propensity matched plots (IC with CAD & CAD without IC)

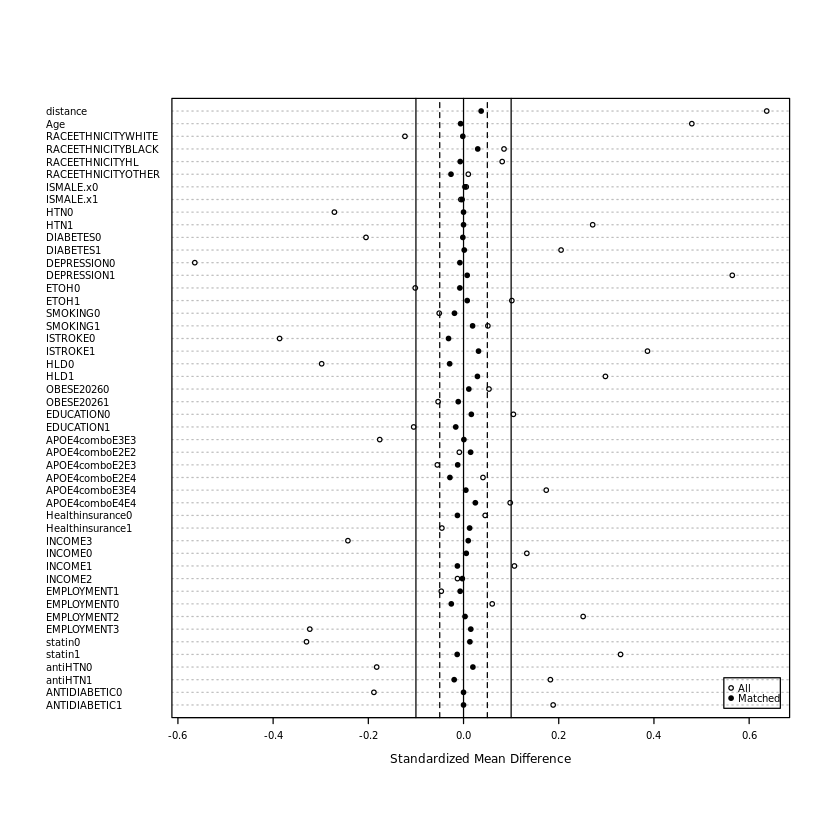

Notes:

ISMALE: Male sex (yes or no); HTN: Hypertension; ETOH: Alcohol use; HLD: Hyperlipidemia; ISTROKE: Ischemic stroke; OBESE2026: BMI ≥ 30; statin: statin drug use; antiHTN: antihypertensive drug use

CAD: Coronary Artery Disease

IC: Impaired Cognitive Disease

| eTable 2: Baseline characteristics in IC vs CAD (AllofUS, v8, Age ≥60)  before and after propensity matching | | | | | | |
| --- | --- | --- | --- | --- | --- | --- |
|  | Before Propensity Matching | | | After Propensity Matching | | |
|  | CAD without IC | IC with CAD | P value | CAD without IC | IC with CAD | P value |
|  | N=21,441 | N=908 |  | N=2,724 | N=908 |  |
| Demographics | | | | | | |
| Age | 73.85 (60-104) | 77.83 (60-100) | p<0.001 | 77.87 (60-104, SD:8.3) | 77.83 (60-100, SD:8.3) | p=0.87 |
| Female Sex | 8583 (40%) | 366 (40.3%) | p=0.867 | 1,094 (40.2%) | 366 (40.3%) | p=0.94 |
| Health insurance | 20,739 (96.7%) | 870 (95.8%) | p=0.133 | 2,603 (95.6%) | 870 (95.8%) | p=0.74 |
| Race | | | | | | |
| White race | 15,032 (70.1%) | 583 (64.2%) | **p<0.001** | 1,751 (64.3%) | 583 (64.2%) | p=1.0 |
| Black race | 2,561 (11.9%) | 136 (15.0%) | **p=0.007** | 379 (13.9%) | 136 (15.0%) | p=0.45 |
| Other | 1,968 (9.2%) | 86 (9.5%) | **p=0.81** | 279 (10.2%) | 86 (9.5%) | p=0.54 |
| Hispanic | 1,880 (9.2%) | 112 (12.3%) | **p=0.008** | 315 (10.2%) | 103 (11.3%) | p=0.9 |
| Annual household income | | | | | | |
| Not reported | 4630 (21.6%) | 250 (27.5%) | **p<0.001** | 743 (27.3%) | 250 (27.5%) | p=0.99 |
| Less than equal to $25,000 | 4169 (19.4%) | 218 (24.0%) | **p<0.001** | 669 (24.6%) | 218 (24.0%) | p=0.99 |
| Between $25,000-$75,000 | 5954 (27.8%) | 247 (27.2%) | p=0.73 | 744 (27.3%) | 247 (27.2%) | p=0.99 |
| More than $75,000 | 6688 (31.2%) | 193 (21.3%) | **p<0.001** | 568 (20.9%) | 193 (21.3%) | p=0.79 |
| Employment | | | | | | |
| Not reported | 340 (1.6%) | 23 (2.5%) | **p=0.04** | 80 (2.9%) | 23 (2.5%) | p=0.60 |
| Unemployed | 3717 (17.3%) | 142 (15.6%) | p=0.20 | 433 (15.9%) | 142 (15.6%) | p=0.98 |
| Retired | 12,237 (57.1%) | 624 (68.7%) | **p<0.001** | 1868 (68.6%) | 624 (68.7%) | p=0.98 |
| Employed | 5,147 (24.0%) | 119 (13.1%) | **p<0.001** | 343 (12.6%) | 119 (13.1%) | p=0.73 |
| Education | | | | | | |
| Highest Education | 10,009 (46.7%) | 377 (41.5%) | **p=0.002** | 1,153 (42.3%) | 377 (41.5%) | p=0.67 |
| Traditional risks | | | | | | |
| Depression | 8,102 (37.8%) | 588 (64.8%) | **p<0.001** | 1,754 (64.4%) | 588 (64.8%) | p=0.84 |
| Ischemic Stroke | 2,491 (11.6%) | 265 (29.2%) | **p<0.001** | 756 (27.8%) | 265 (29.2%) | p=0.40 |
| Diabetes | 10,057 (46.9%) | 518 (57.0%) | **p<0.001** | 1552 (57.0%) | 518 (57.0%) | p=0.97 |
| Hyperlipidemia | 19,805 (92.4%) | 883 (97.2%) | **p<0.001** | 2,636 (96.8%) | 883 (97.2%) | p=0.47 |
| Hypertension | 19,379 (90.4%) | 870 (95.8%) | **p<0.001** | 2,610 (95.8%) | 870 (95.8%) | p=1.0 |
| Obesity | 9,729 (45.4%) | 388 (42.7%) | p=0.117 | 1,179 (43.3%) | 388 (42.7%) | p=0.77 |
| Smoking | 5,299 (24.7%) | 245 (27.0%) | p=0.121 | 712 (26.1%) | 245 (27.0%) | p=0.62 |
| Alcohol Use | 1,992 (9.3%) | 115 (12.7%) | **p<0.001** | 338 (12.4%) | 115 (12.7%) | p=0.84 |
| Statin use | 16,438 (76.7%) | 795 (87.6%) | **p<0.001** | 2,397 (88.0%) | 795 (87.6%) | p=0.72 |
| Antihypertensive use | 18,503 (86.3%) | 830 (91.4%) | **p<0.001** | 2,505 (92.0%) | 830 (91.4%) | p=0.600 |
| Antidiabetic | 10,161 (47.4%) | 515 (56.7%) | **p<0.001** | 1545 (56.7%) | 515 (56.7%) | p=1.00 |
| APO (ε) genotypes | | | | | | |
| APOε2ε2 | 132 (0.6%) | 5 (0.6%) | p=0.97 | 12 (0.4%) | 5 (0.6%) | p=1.0 |
| APOε2ε3 | 2,478 (11.6%) | 90 (9.9%) | p=0.14 | 280 (10.3%) | 90 (9.9%) | p=0.80 |
| APOε3ε3 | 13,312 (62.1%) | 484 (53.3%) | **p<0.001** | 1,451 (53.3%) | 484 (53.3%) | p=0.9 |
| APOε2ε4 | 468 (2.2%) | 26 (2.9%) | p=0.21 | 91 (3.3%) | 26 (2.9%) | p=0.82 |
| APOε3ε4 | 4,629 (21.6%) | 268 (29.5%) | **p<0.001** | 798 (29.3%) | 268 (29.5%) | p=0.93 |
| APOε4ε4 | 422 (2.0%) | 35 (3.9%) | **p<0.001** | 92 (3.4%) | 35 (3.9%) | p=0.56 |
| *P value <0.05 (Significance in bold) | | | | | | |
| IC: Impaired Cognitive Disease | | | | | | |
| CAD: Coronary Artery Disease | | | | | | |
